# Immune Changes in Pregnancy: Associations with Pre-existing Conditions and Obstetrical Complications at the 20th Gestational Week - A Prospective Cohort Study

**DOI:** 10.1101/2023.08.10.23293934

**Authors:** David Westergaard, Agnete Troen Lundgaard, Kilian Vomstein, Line Fich, Kathrine Vauvert Römmelmayer Hviid, Pia Egerup, Ann-Marie Hellerung Christiansen, Josefine Reinhardt Nielsen, Johanna Lindman, Peter Christoffer Holm, Tanja Schlaikjær Hartwig, Finn Stener Jørgensen, Anne Zedeler, Astrid Marie Kolte, Henrik Westh, Henrik Løvendahl Jørgensen, Nina la Cour Freiesleben, Karina Banasik, Søren Brunak, Henriette Svarre Nielsen

## Abstract

**Background:** Pregnancy is a complex biological process and serious complications can arise when the delicate balance between the maternal immune system and the semi-allogeneic fetal immune system is disrupted or challenged. Gestational diabetes mellitus (GDM), pre-eclampsia, preterm birth, and low birth weight, pose serious threats to maternal and fetal health. Identification of early biomarkers through an in-depth understanding of molecular mechanisms is critical for early intervention.

**Methods:** We analyzed the associations between 47 proteins involved in inflammation, chemotaxis, angiogenesis, and immune system regulation, maternal and neonatal health outcomes, and the baseline characteristics and pre-existing conditions (diseases and obstetric history) of the mother in a prospective cohort of 1,049 pregnant women around the 20th gestational week. Bayesian linear regression models were used to examine the impact of risk factors on biomarker levels and Bayesian cause-specific parametric proportional hazards models were used to analyze the effect of biomarkers on maternal and neonatal health outcomes. Finally, we evaluated the predictive value of baseline characteristics and the 47 proteins using machine-learning models. Shapley additive explanation (SHAP) scores were used to dissect the machine learning models to identify biomarkers most important for predictions.

**Results:** Associations were identified between specific inflammatory markers and existing conditions, including maternal age and pre-pregnancy BMI, chronic diseases, complications from prior pregnancies, and COVID-19 exposure. Smoking during pregnancy significantly affected GM-CSF and 9 other biomarkers. Distinct biomarker patterns were observed for different ethnicities. In obstetric complications, IL-6 inversely correlated with pre-eclampsia risk, while acute cesarean section and birth weight to gestational age ratio were linked to markers such as VEGF or PlGF. GDM was associated with IL-1RA, IL-17D, and Eotaxin-3. Severe PPH correlated with CRP and proteins of the IL-17 family. Predictive modeling using MSD biomarkers yielded ROC-AUC values of 0.708 and 0.672 for GDM and pre-eclampsia, respectively. Significant predictive biomarkers for GDM included IL-1RA and Eotaxin-3, while pre-eclampsia prediction yielded highest predictions when including MIP-1β, IL-1RA, and IL-12p70.

**Conclusion:** Our study provides novel insights into the interplay between preexisting conditions and immune dysregulation in pregnancy. These findings contribute to our understanding of the pathophysiology of obstetric complications and the identification of novel biomarkers for early intervention(s) to improve maternal and fetal health.

## Introduction

During pregnancy there is a complex interaction between the maternal immune system and the semi-allogeneic fetus^1^. The maternal immune system is highly regulated throughout pregnancy, and successful pregnancy requires a balance between tolerance and suppression. Obstetric complications are common, affecting more than one in four pregnancies, and immune dysregulation is involved in the pathogenesis of a range of complications, including gestational diabetes mellitus (GDM), pre-eclampsia, preterm birth, and low birth weight^2,3^. Immune dysregulation may be due to preexisting or subclinical diseases, complications from prior pregnancies, or viral infections. Indeed, during pregnancy, the maternal immune system seems to be more challenged by viral infections such as influenza^4,5^, respiratory syncytial virus (RSV)^6^, severe acute respiratory syndrome (SARS-CoV)^7,8^, and Middle East Respiratory Syndrome (MERS-CoV)^9^. Nonetheless, many pregnancy complications arise without prior known risk factors. Consequently, identifying biomarkers that are present early in pregnancy prior to the manifestation of the pathology is crucial to enable timely action and improve maternal and fetal health outcomes.

Pregnancy complications such as preterm birth and early onset pre-eclampsia increase the risk in subsequent pregnancies. Conversely, a prior uncomplicated pregnancy decreases risk of complications in future pregnancies, possibly due to enduring immune alterations favoring fetal tolerance^10^. An important aspect of the immune alteration in pregnancy involves the balance of pro- and anti-inflammatory responses; typically, the immune system maintains a balance between T helper-1 (Th-1) cells, associated with cell-mediated immunity and inflammation, and T helper-2 (Th-2) cells, supporting humoral immunity and linked with anti-inflammatory responses^11,12^. During pregnancy, the systemic maternal immune response shifts towards a more anti-inflammatory state which promotes fetal tolerance. Disruptions in this balance, particularly a switch towards a pro-inflammatory state, can lead to pregnancy complications such as pregnancy loss, pre-eclampsia, and preterm birth^11^. Even though it’s important to understand these risks, we don’t know much about how previous pregnancies affect certain biomarker levels related to immune regulation in the current pregnancy. However, promising research highlights the potential for early detection of preterm birth using cell-free RNA or a protein panel, as well as pre-eclampsia through a combination of sFlt-1/PlGF and ultrasound, among other methods^13–15^. A deeper understanding of the impact of previous pregnancies and the biological changes that occur before labor starts is crucial. It serves as the foundation for the development of diagnostic tests and tools aimed at improving maternal and fetal health outcomes.

Overall, the link between immune adaptation during pregnancy and obstetric complications is complex and multifaceted. While immune changes are necessary for a successful pregnancy and fetal development, they can also increase the risk of obstetric complications. Prophylactic treatment is an option for many obstetric complications, such as low-dose aspirin or progesterone. However, this requires precise risk stratification of pregnant women. Early biomarkers of later complications facilitate early interventions that can improve fetal and maternal health.

Here, we present results that build towards a deeper understanding of the immune system, the interplay between diseases and obstetric history and the immune system and its association with obstetric complications in 1,049 pregnant women

## Methods

### Ethical approvals

The PREGCO study was approved by the Knowledge Centre for Data Protection and Compliance, The Capital Region of Denmark (P-2020-255), and by the Scientific Ethics Committee of the Capital Region of Denmark (journal number H-20022647). All PREGCO participants provided written informed consent.

Danish legislation allows for register-based research to be conducted without the consent of participants and without ethical committee approval. Registry data, i.e. the Danish Medical Birth Registry, was held at Statistics Denmark, which is the Danish national statistical institution.

### Study design and participants

PREGCO is a prospective cohort of pregnant women from Copenhagen University Hospital, Hvidovre, Denmark, that took place during the first pandemic wave in Denmark, between 4th April 2020 and 3rd July 2020^16,17^. Participants were invited to participate at the second trimester malformation scan (gestational weeks 18-22). All pregnant women in Denmark are offered this scan and more than 90% accept. Copenhagen University Hospital Hvidovre serves approximately 12% of pregnant women in Denmark (∼7,200 births/year). Participants filled out a questionnaire including, but not limited to, pre-pregnancy body mass index (BMI), smoking, prior pregnancy outcomes and complications, and pre-existing chronic conditions. Baseline characteristics and follow-up information were obtained from the electronic health record, available throughout the whole study period. This included maternal age, gestational age at birth, sex of the child, multiple pregnancy, birth weight, results from the combined first trimester screening examination (e.g., nuchal fold thickness, pregnancy-associated plasma protein A (PAPP-A), β-human choriogonadotropin (βhCG), and crown-rump length), second trimester malformation scan (e.g., head circumference and femur length), obstetric complications (e.g., GDM, pre-eclampsia, acute cesarean section), and mode of delivery (spontaneous delivery, induction of labor, or cesarean section). See Supplementary Table 5 for a complete list. Furthermore, serum was collected at the 12th and 20th gestational week scans to screen for the presence of SARS-CoV-2 antibodies (IgG and IgM) using the iFlash 1800 assay^16,17^. Serum samples from the second trimester malformation scan were also used to measure a panel of 47 inflammatory markers (described below). The study was completed and all pregnancies ended before the approval and introduction of SARS-CoV-2 vaccines in Denmark (27th December 2020).

### Comparison with the general population

The Danish Medical Birth Register (DMBR)^18^ was used to compare the PREGCO cohort with all births in Denmark during the same period. DMBR, established in 1973, includes detailed data on all births in Denmark and primarily comprises data from the Danish National Patient Registry supplemented with information on pre-pregnancy BMI and smoking in the first trimester collected at the combined first trimester screening examination. We compared maternal age, pre-pregnancy BMI, smoking, parity, number of pregnancy losses, and sex of the child.

### Meso Scale Diagnostics inflammatory markers

The setup for measuring the inflammatory markers has been described by Kjerulff *et al*^19^. Inflammatory markers from the second trimester malformation scan serum sample were measured using the Meso Scale Diagnostics (MSD) V-PLEX Human Biomarker 54-Plex kit, measuring 47 biomarkers involved in inflammation, chemotaxis, angiogenesis, and immune system regulation (due to poor quality observed for the TH-17 panel^19^, the panel was excluded). The samples were spread across sixteen 96-well plates. We performed preprocessing and median normalization of measured intensities to minimize batch variation, as described in detail in the Supplementary Text (Supplementary Figure 1).

### Exposures and outcomes

We investigated the influence of pre-existing conditions (diseases and obstetric history) and diseases on biomarker levels at the second trimester malformation scan. This included smoking during pregnancy, number of prior live births, number of prior pregnancy losses, polycystic ovary syndrome (PCOS), endometriosis, inflammatory bowel disease, GDM in the current or previous pregnancy, pre-eclampsia in a prior pregnancy, vaginal bleeding in early pregnancy, use of assisted reproductive technology (ART, divided into intrauterine insemination (IUI) and in-vitro fertilization (IVF)), and Coronavirus disease 2019 (COVID-19) infection in the current pregnancy. The COVID-19 assay and cut-offs have been previously described by Freiesleben *et al* and Egerup *et al*^16,17^. Second, we also investigated the association between inflammatory markers and common obstetric complications, namely GDM, pre-eclampsia, gestational duration, acute cesarean section, and the ratio of birth weight to gestational duration (Supplementary Table 1). To reduce redundancy in the statistical analysis, we identified highly correlated markers using the Hobohm II algorithm^20^. We tested multiple thresholds and found that a Spearman correlation cut-off of 0.5 sufficiently removed redundant markers (Supplementary Figure 2). For further details, see Supplementary Information.

### Machine learning models to predict obstetrical complications

We evaluated the predictive value of the clinical measures and the MSD biomarkers using two machine learning models, namely a logistic regression model with an L1 penalty and LightGBM. Missing values were imputed using the mean and mode for continuous and categorical variables, respectively, and scaled for the logistic regression model. Imputation and scaling were done strictly on training data. LightGBM is a gradient-boosted model that natively handles missing values and categorical data, and scaling is not needed. As features we used the 47 Meso Scale Diagnostics inflammatory markers, age, BMI, previous live births, fetal abdominal circumference, fetal abdominal diameter, fetal head circumference, fetal femur length, PAPP-A MoM, βhCG MoM, and the difference between gestational age measured from crown-rump length and last menstruation at the combined first trimester screening examination.

The models’ generalizability was evaluated using a nested cross-validation (CV) procedure. Here, the Nested CV involved an outer loop for model evaluation (interval validation) and an inner loop for hyperparameter optimization (development data). The outer loop was a five-fold stratified CV, and the inner loop was a five-fold stratified CV repeated five times. Hyperparameters were optimized using the Optuna^21^ framework with the Tree-structured Parzen Estimator algorithm. Supplementary Table 2 provides the ranges of hyperparameters that were explored. A total of 500 hyperparameter combinations were evaluated across the inner CV loops to identify the most effective configuration for the model. The models were optimized to minimize the binary cross entropy (BCE). We selected the model with the lowest average inner CV BCE. We evaluated the Area Under the Receiver Operating Characteristic Curve (ROC-AUC) and the area under the precision-recall curve (AUPRC) by averaging the scores from the outer CV. Feature importance was evaluated using Shapley Additive Explanations (SHAP) on the hold-out outer fold. The ROC-AUC ranges from 0.5 (random) to 1.0 (perfect). However, in cases of severe class imbalance, the ROC-AUC may be biased. Therefore, we also evaluated the AUPRC. The AUPRC ranges from 0 to 1. However, the baseline value (corresponding to a random classifier) is equivalent to the prevalence. 95% Confidence intervals for the ROC-AUC and AUPRC were calculated using a bootstrap approach, with 1,000 repetitions. We used the linear (for logistic regression) or tree (for LightGBM) explainer algorithm. Results were visualized as the mean absolute SHAP values. Machine learning pipelines were implemented as a snakemake workflow, using elements from Optuna, scikit-learn, and SHAP.^21–23^

### Statistical analysis

Baseline characteristics were summarized as mean (standard deviation, SD) or median (interquartile range, IQR), where appropriate. Baseline characteristics were compared to data from the DMBR for all births in 2020 using a Z-test or χ^2^ test. Heatmaps were created by calculating pairwise Spearman’s correlation coefficients. Clustering was done using hierarchical complete clustering. Only MSD biomarkers were used for clustering.

The association between prior existing conditions and inflammatory marker levels and Birth weight-Gestational Age ratio was estimated using a Bayesian robust linear regression model to accommodate outliers^24^.

The association between inflammatory markers and GDM, pre-eclampsia, gestational duration, acute cesarean section, severe postpartum hemorrhage (PPH), and any complication was estimated using a cause-specific Bayesian time-to-event model, in which the baseline hazard was modeled using an M-spline^25^. Women lost to follow-up were censored at their last contact with the hospital. All analyses were adjusted for age, pre-pregnancy BMI, and gestational age at enrollment. The association between prior conditions and inflammatory markers was further adjusted for the ultrasound estimated gestational age at the first trimester risk assesment scan. Obstetric outcomes were adjusted for outcome-specific variables that were identified based on expert and literature review. Missing values were imputed using multiple imputation as implemented in MICE^26^, with predictive mean matching. Chains were run for 100 iterations and five imputation data sets were created. Each imputed data set was analyzed separately in the Bayesian models and the posteriors were then combined. Conservative prior distributions were used for the Bayesian models, providing a regularizing effect. Estimates are reported as the median and the 95% credible interval (bCI) unless otherwise stated. See Supplementary Methods for a more detailed description of the methods and models. All models were fitted using rstanarm^25^ or brms^24^.

## Results

### Cohort characteristics

A total of 1,064 women were enrolled in this study during the second trimester malformation scan at 20 weeks. The cohort constitutes 75% of all pregnant women asked to participate (flowchart shown in Figure 1A). Of these, 15 were not included due to multiple pregnancy and 18 were lost to follow-up due to a change of hospital not using the EPIC electronic platform for medical records or home delivery. The baseline characteristics for participants are shown in Table 1. Compared to all births in Denmark in 2020, maternal age, pre-pregnancy BMI, and the number of previous live births are within the expected range, albeit they are on average 1 year older, 0.7 BMI units lower, and have a higher frequency of primipara and prior pregnancy losses (Supplementary Table 3). 33 women (3.1%) had elevated SARS-CoV-2 IgG antibody levels indicating COVID-19 infection prior to the second trimester malformation scan. During the study, 369 (34.7%) women experienced at least one of the complications severe PPH (12.2%), preterm birth (11%), GDM (8.2%), pre-eclampsia (4.3%), or acute cesarean section (11.8%) after enrollment (Table 2).

**Figure 1:**
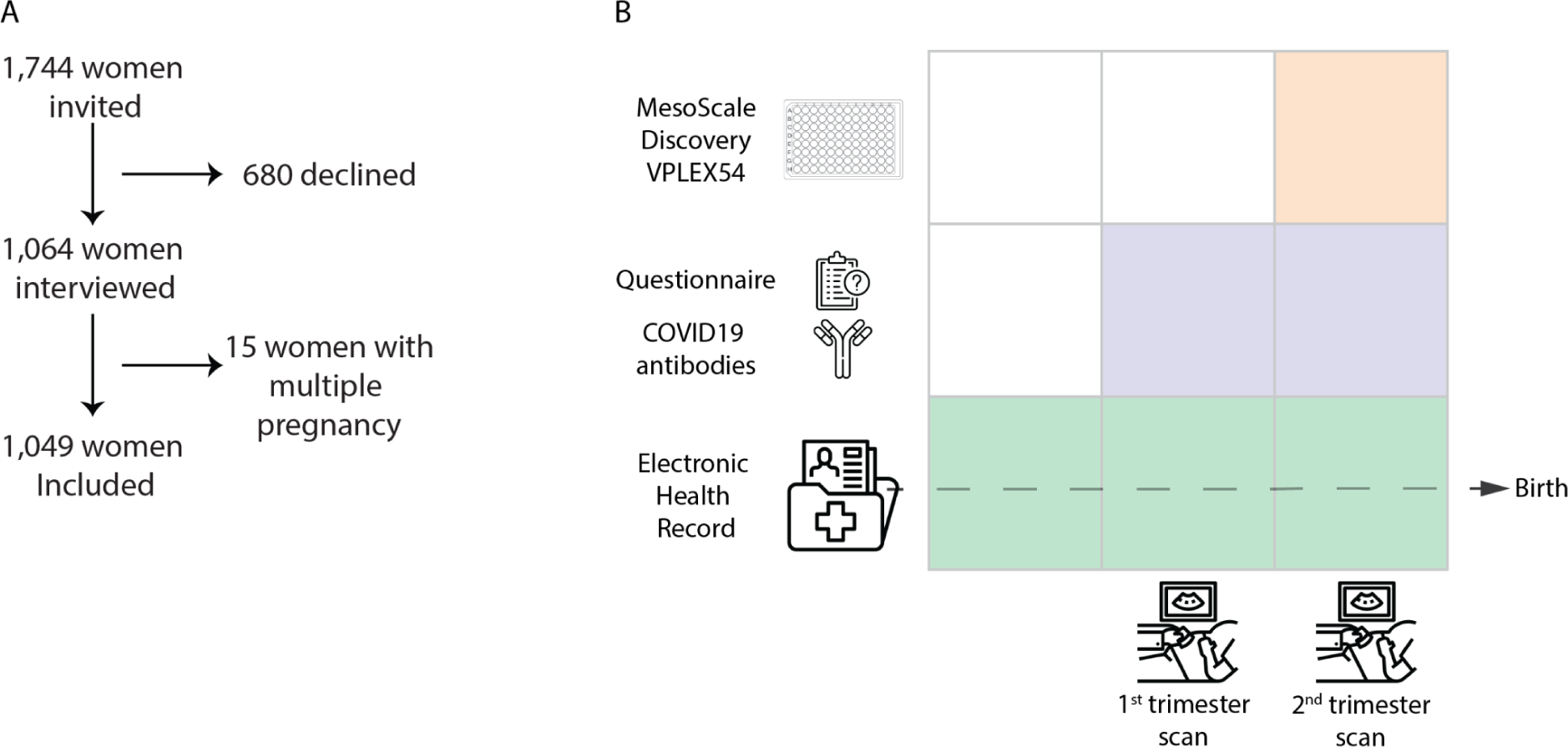
(a) Flowchart for inclusion, (b) Timepoints for data collection. Data from the electronic health record was obtained throughout the pregnancy and postpartum.

**Table 1:**
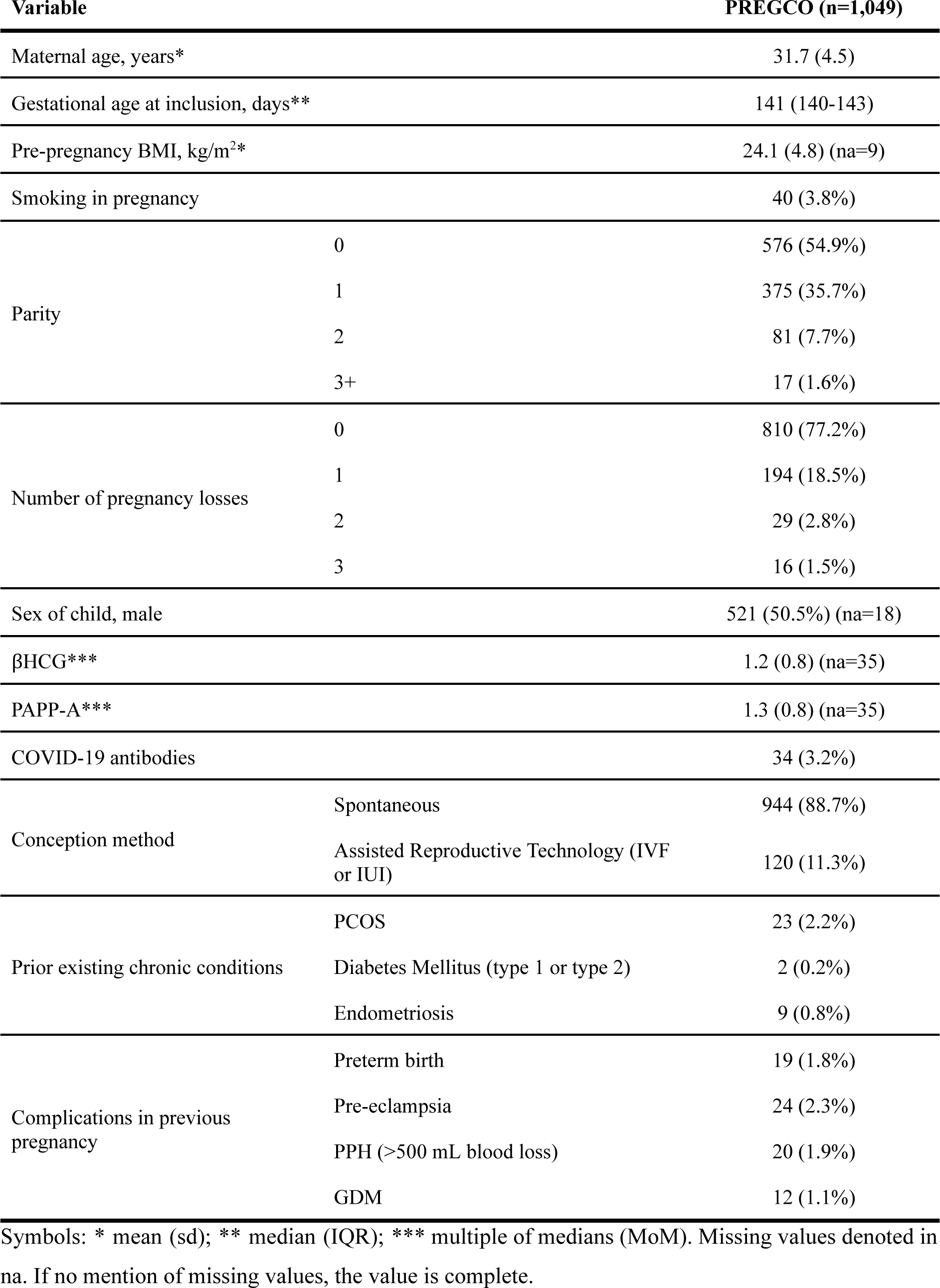
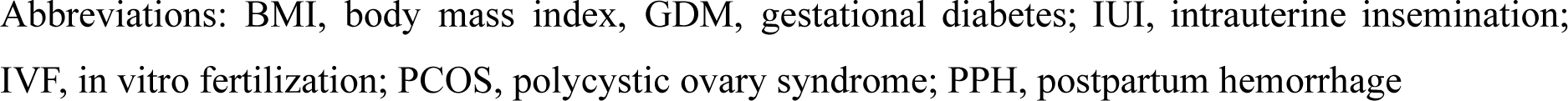
Baseline characteristics for the 1,049 women included in the PREGCO cohort.

| Variable |  | PREGCO (n=1,049) |
| --- | --- | --- |
| Maternal age, years* |  | 31.7 (4.5) |
| Gestational age at inclusion, days** |  | 141 (140-143) |
| Pre-pregnancy BMI, kg/m<sup>2</sup>* |  | 24.1 (4.8) (na=9) |
| Smoking in pregnancy |  | 40 (3.8%) |
| Parity | 0 | 576 (54.9%) |
|  | 1 | 375 (35.7%) |
|  | 2 | 81 (7.7%) |
|  | 3+ | 17 (1.6%) |
| Number of pregnancy losses | 0 | 810 (77.2%) |
|  | 1 | 194 (18.5%) |
|  | 2 | 29 (2.8%) |
|  | 3 | 16 (1.5%) |
| Sex of child, male |  | 521 (50.5%) (na=18) |
| βHCG*** |  | 1.2 (0.8) (na=35) |
| PAPP-A*** |  | 1.3 (0.8) (na=35) |
| COVID-19 antibodies |  | 34 (3.2%) |
| Conception method | Spontaneous | 944 (88.7%) |
|  | Assisted Reproductive Technology (IVF or IUI) | 120 (11.3%) |
| Prior existing chronic conditions | PCOS | 23 (2.2%) |
|  | Diabetes Mellitus (type 1 or type 2) | 2 (0.2%) |
|  | Endometriosis | 9 (0.8%) |
| Complications in previous pregnancy | Preterm birth | 19 (1.8%) |
|  | Pre-eclampsia | 24 (2.3%) |
|  | PPH (>500 mL blood loss) | 20 (1.9%) |
|  | GDM | 12 (1.1%) |

Symbols: \* mean (sd); \*\* median (IQR); \*\*\* multiple of medians (MoM). Missing values denoted in na. If no mention of missing values, the value is complete.
Abbreviations: BMI, body mass index, GDM, gestational diabetes; IUI, intrauterine insemination; IVF, in vitro fertilization; PCOS, polycystic ovary syndrome; PPH, postpartum hemorrhage

**Table 2:** Outcome characteristics.

| Variable |  | PREGCO |
| --- | --- | --- |
| Birth weight (g)* |  | 3499 (554) |
| Gestational age (days)** |  | 280 (273; 287) |
| Preterm (<37+0) |  | 116 (11%) |
| Birth weight / Gestational age** |  | 12.6 (1.8) |
| Severe postpartum hemorrhage (PPH) |  | 130 (12.2%) |
| Pre-eclampsia |  | 46 (4.3%) |
| Gestational diabetes mellitus (GDM) |  | 87 (8.2%) |
| Mode of delivery | Spontaneous vaginal birth | 634 (60.7%) |
|  | Acute cesarean section | 126 (11.8%) |
|  | Elective cesarean section | 95 (8.9%) |
|  | Induced birth | 235 (22.1%) |
| Any complication | (Severe PPH, preterm birth, GDM, pre-eclampsia, acute cesarean section) | 369 (34.7%) |

Symbols: \* *mean (sd)*; \*\* *median (IQR)*.
Abbreviations: GDM, gestational diabetes mellitus; PPH, postpartum hemorrhage

### Biomarker profile clustering

Clustering analysis did not reveal any strong relationships between the women’s inflammatory profiles and their prior conditions, diseases, or later complications (Figure 2, Supplementary Figure 3a). When we examined the pairwise correlations between markers, we found that they did not necessarily cluster by panel or marker group (Supplementary Figure 3b). Instead, we observed one large cluster and some minor clusters. The largest cluster, with the strongest correlations, consisted of MCP-4, TARC, IL-8, VEGF-C, and IL-7. MCP-4, TARC, IL-8, VEGF-C, and IL-7 are all cytokines linked to various inflammatory diseases.

**Figure 2:**
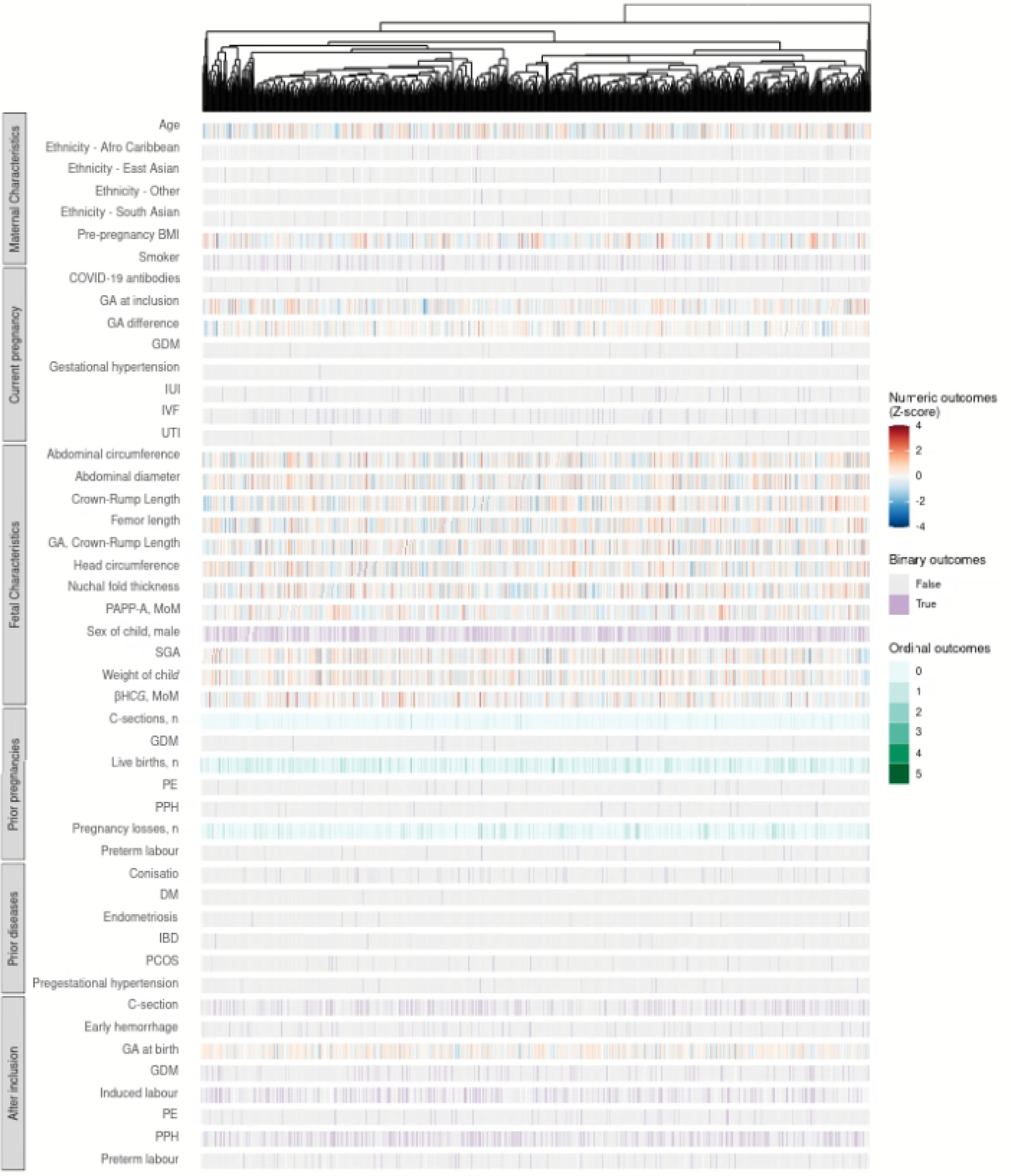
Heatmap of phenotypes across participants using hierarchical clustering based on MSD biomarkers. Phenotypes are divided into maternal characteristics, pregnancy-related outcomes for the current pregnancy (observed before or at inclusion), fetal characteristics, pregnancy-related outcomes for prior pregnancies, prior diseases to the current pregnancy, and phenotypes observed after inclusion. BMI, body mass index; C-section, cesarean section; DM, diabetes mellitus; GA, gestational age; GDM, gestational diabetes mellitus; IBD, irritable bowel diseases; IUI, intrauterine insemination; IVF, in vitro fertilization; MoM, multiples of medians; PCOS, polycystic ovary syndrome; PE, Pre-eclampsia; PPH, postpartum hemorrhage; SGA, small for gestational age; UTI, urinary tract infection

The strongest anti-correlation observed was between Flt-1 and VEGF (-0.58, 95% CI -0.62; -0.53, Pearson correlation). By binding VEGF and blocking the membrane-bound receptors, soluble Flt-1 functions as a naturally occurring antagonist of VEGF. Soluble Flt-1 also binds placental growth factor (PlGF) that plays a crucial role in the growth and development of blood vessels, particularly during pregnancy and fetal development^27^. Dysregulation of the interaction between Flt-1 and VEGF has been linked to various diseases and disorders, including cancer and cardiovascular disease.

### Pre-existing conditions, obstetrical history, and maternal characteristics

A number of previously existing conditions correlated to the measured inflammatory markers. However, there was no widespread pleiotropy, i.e., each exposure had its own biomarker signature (Figure 3). A majority of biomarkers were affected by maternal age (15/47) and pre-pregnancy BMI (26/47). Conditions altering expression levels included chronic diseases (endometriosis, PCOS), complications in prior pregnancies (pre-eclampsia, GDM, preterm birth), and COVID-19. COVID-19 was associated with a change in expression of two inflammatory markers, IFN-γ and IL-13. Smoking in the current pregnancy had a profound effect on ten biomarkers, including GM-CSF (β=0.05, 95% bCI 0.02; 0.09). GM-CSF is a known regulator of fetal growth and the association between smoking and increased GM-CSF levels is hypothesized to be through an activation of the EGFR signaling pathway^28^. GM-CSF was also upregulated in pregnancies with a female child, albeit to a lower degree (β=0.02, 95% bCI 0.01; 0.03). Altered CRP levels were associated with complications occurring in prior pregnancies, e.g. pre-eclampsia (β=-0.45, 95% bCI -0.89; -0.03) and preterm birth (β=0.76, 95% bCI 0.28; 1.3). The effect of pre-eclampsia and preterm birth is most likely medically induced due to preventive treatment with aspirin and progesterone, respectively. The number of previous live births was also associated with lower CRP levels, which could explain part of the mechanism behind the reduced risk of pre-eclampsia (β=0.16, 95% bCI 0.06; 0.27).

**Figure 3:**
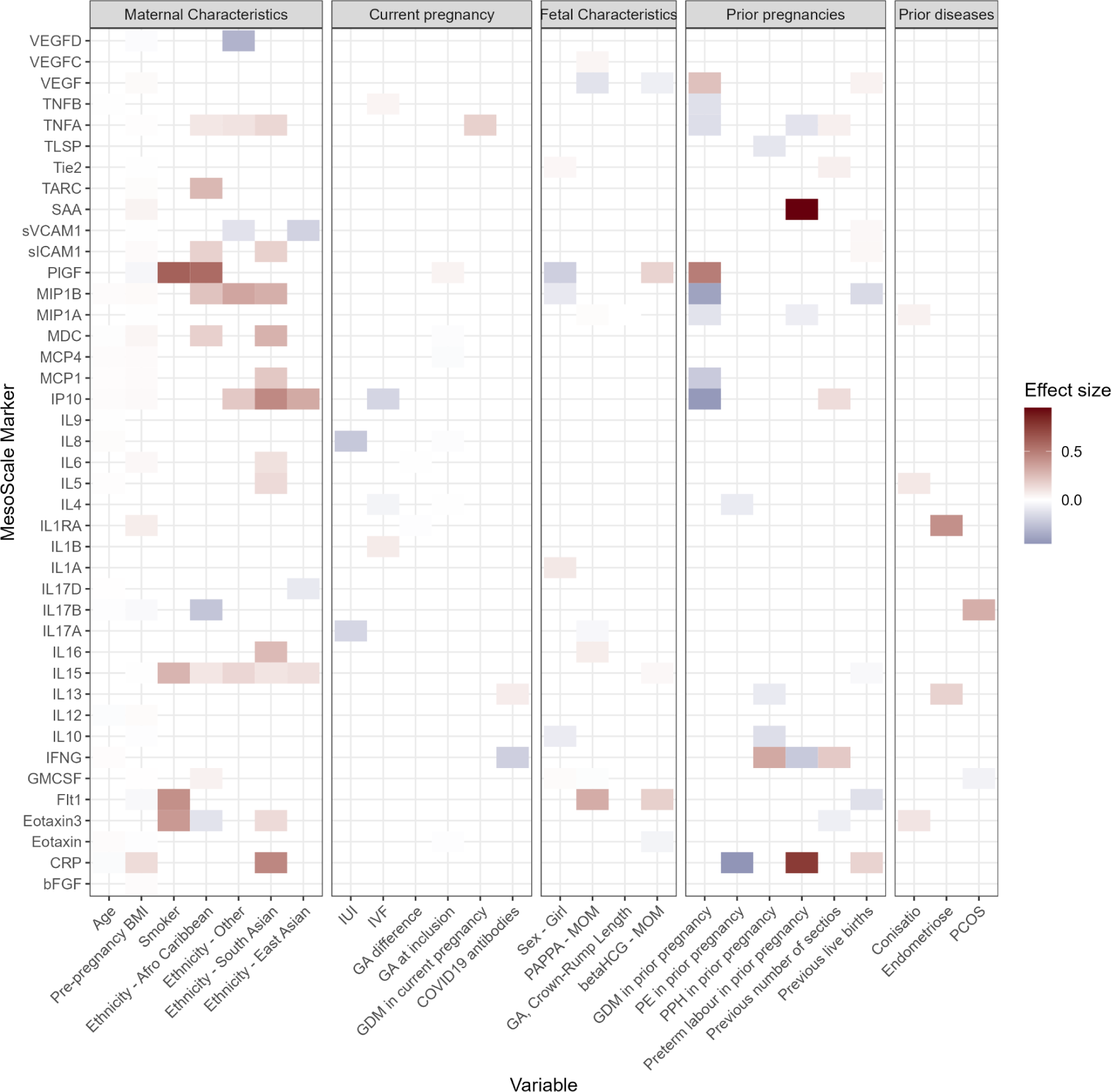
Previous pregnancies and pre-existing conditions effects on inflammatory markers. Effect size is increase or decrease in standard deviations. Only associations where the 95% Bayesian Credible Interval does not include zero are shown. BMI, body mass index; GA, gestational age; GDM, gestational diabetes mellitus; IUI, intrauterine insemination; IVF, in vitro fertilization; MOM, multiples of medians; PCOS, polycystic ovary syndrome; PE, pre-eclampsia; PPH, postpartum hemorrhage

TNF-α levels were increased in women with GDM prior to the second trimester malformation scan (β =0.17, 95% bCI 0.01; 0.34) and decreased in women with GDM in prior pregnancies (β=-0.14, 95% bCI -0.27; -0.01). TNF-α is a marker of insulin resistance in pregnancy^29^ and our findings indicate that insulin resistance is not affected in the longer term in GDM pregnancies. This suggests that increased TNF-α levels can be attributed to the acute pathogenic process.

Ethnicity had a major influence on the variation of inflammatory marker levels, such as CRP, Eotaxin-3, IL-15, IL-16, IP-10, and VEGF-D, amongst others (Figure 3). These findings highlight the need for more research on the role of ethnicity in pregnancy complications and the underlying mechanisms. This research could inform the development of personalized interventions to reduce the risk of adverse pregnancy outcomes by taking into account reference ranges may be specific to ethnic groups.

### Later obstetric complications

Following similarity reduction, we estimated the association between 41 markers and 11 different outcomes (Figure 4). We found a number of unique markers that were associated with each condition. Increasing levels of IL-6 decreased the risk of pre-eclampsia (hazard rate (HR)=0.59, 95% bCI 0.31; 0.97). Prior evidence clearly indicates an association, but the direction of effect is less clear^30,31^. Acute cesarean section had four associated markers (IL-4, IL-5, MDC, MIP-1β) none of which were associated with any of the other adverse outcomes. For the birth weight to gestational age ratio, we found five associated markers: bFGF, GM-CSF, PlGF, sICAM-1, and VEGF. The most profound effect on birth weight to gestational age ratio was seen for vaginal bleeding in early pregnancy, which led to an estimated decrease in percentile of -0.16 (95% bCI -0.06; -0.25). This means that, a woman experiencing early hemorrhage would be expected to be in the 34th percentile (95% bCI 25; 44). In comparison, PlGF, the inflammatory marker with the largest effect, was associated with an increased percentile of 0.04 (95% bCI 0.01; 0.07) per standard deviation.

**Figure 4:**
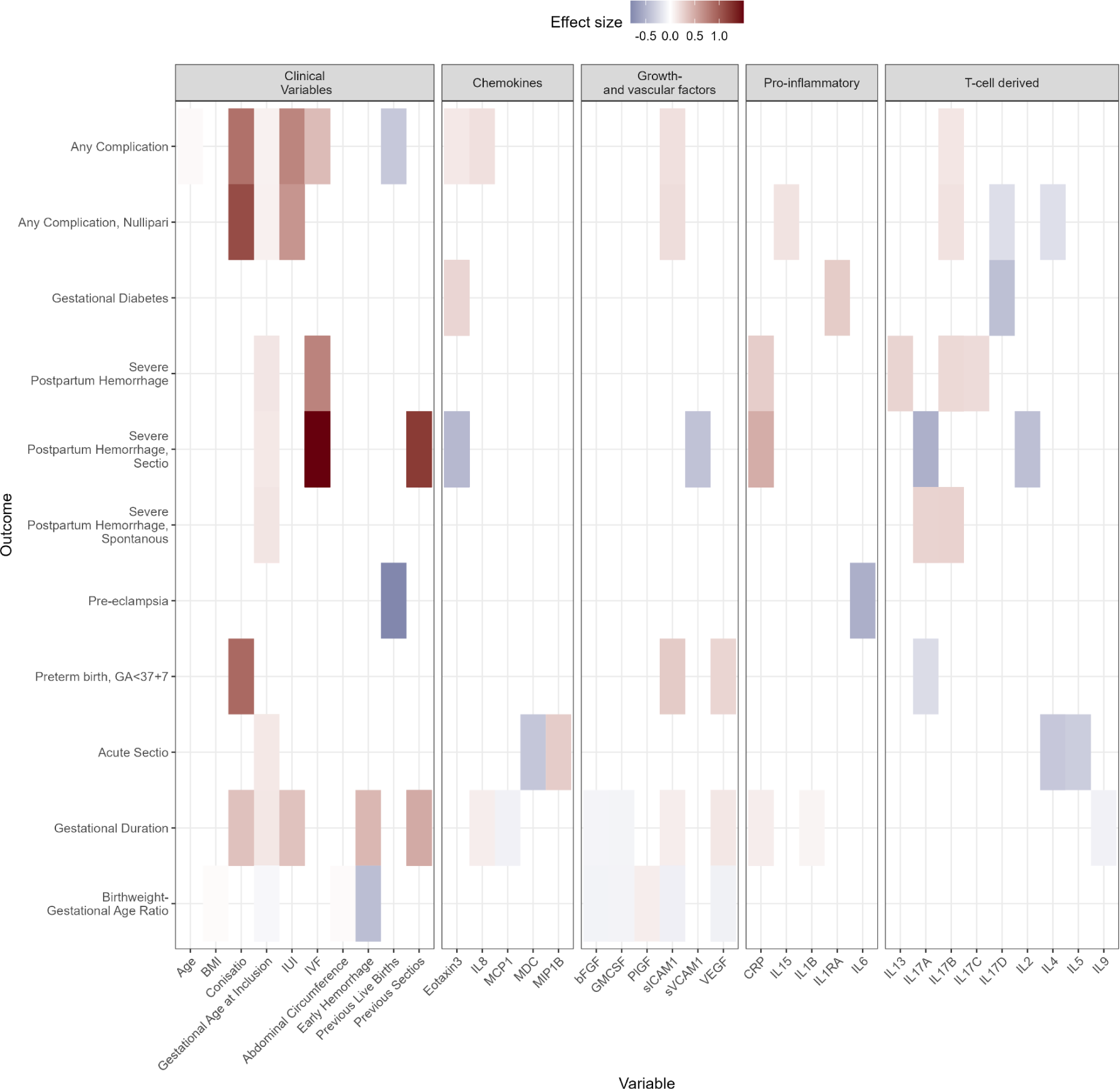
Associations between markers and later obstetrical outcomes. The effect size is the log-hazard rate for all outcomes, except Birth Weight-Gestational Age Ratio (BWGA). The effect of biomarkers on BWGA is a change in percentile from the 50th percentile. Only associations where the 95% Bayesian Credible Interval does not include zero are shown. BMI, body mass index; GA, gestational age; IUI, intrauterine insemination; IVF, in vitro fertilization

GDM was associated with IL-1RA (HR=1.35, 95% bCI 1.04; 1.8), IL-17D (HR=0.66, 95% bCI 0.49; 0.89), and Eotaxin-3 (HR=1.27, 95% bCI 1.11; 1.43). IL-1RA has previously been shown to associate with GDM and has been suggested as a biomarker useful for diagnosing GDM as a complement to blood glucose measurements, as well as to identify GDM patients who are at risk of developing postpartum diabetes^32^. IL-17D is associated with incident type 2 diabetes and progression from normoglycemia to type 2 diabetes^33^. GDM is associated with several chemokines in the protein family where Eotaxin-3/CCL26 belongs^34,35^.

CRP (HR=1.34, 95% bCI 1.04; 1.74), IL-13 (HR=1.29, 95% bCI 1.06; 1.54), IL-17B (HR=1.25, 95% bCI 1.02; 1.53), and IL-17C (HR=1.23, 95% bCI 1.01, 1.49) were all associated with severe PPH. However, none of the effects had a magnitude similar to in vitro fertilization (IVF, HR=2.10, 95% bCI 1.18; 3.66).

### Prediction of obstetric complications

We evaluated the prognostic potential of clinical characteristics combined with MSD biomarkers in predicting five pregnancy-related conditions. The performance of our model was assessed using ROC-AUC values on the development data (inner CV) ranging from 0.610 to 0.697, and similar values were obtained in the interval validation (outer CV), ranging from 0.584 to 0.715. These results indicate that our model generalizes well to new data. The AUPRC values were consistently better than random guessing, as shown in Supplementary Table 6.

Among the pregnancy-related conditions analyzed, pre-eclampsia, GDM and PPH were predicted most accurately using the MSD biomarkers. Specifically, GDM and pre-eclampsia displayed ROC-AUC values of 0.708 (95% CI: 0.644-0.766) and 0.672 (95% CI: 0.580-0.758), respectively, with corresponding AUPRC values of 0.176 (95% CI: 0.126-0.238) and 0.073 (95% CI: 0.042-0.115).

The SHAP analysis further confirmed the classification of IL-1RA and Eotaxin-3 as potential predictive biomarkers for GDM (Figure 6A). Additionally, the analysis highlighted the significance of IL-6 and CRP, proinflammatory markers that were initially overlooked in the preliminary association analysis but have been previously suggested as predictive for GDM^36–38^.

**Figure 5:**
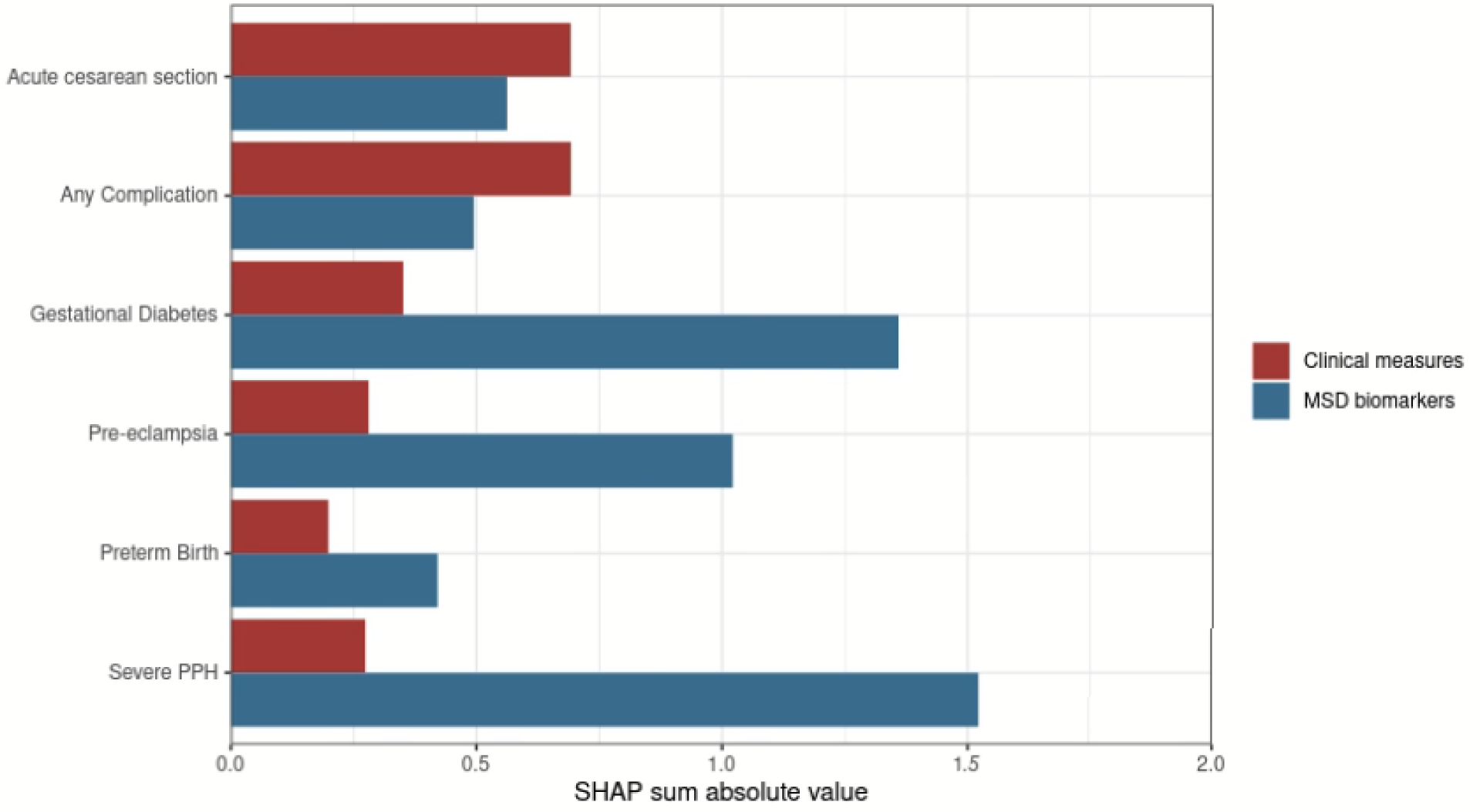
Summarised SHAP values for each feature category (red = clinical measure, blue *= MSD biomarker) across all outcomes investigated in the prognostic model*. PPH, postpartum hemorrhage

**Figure 6:**
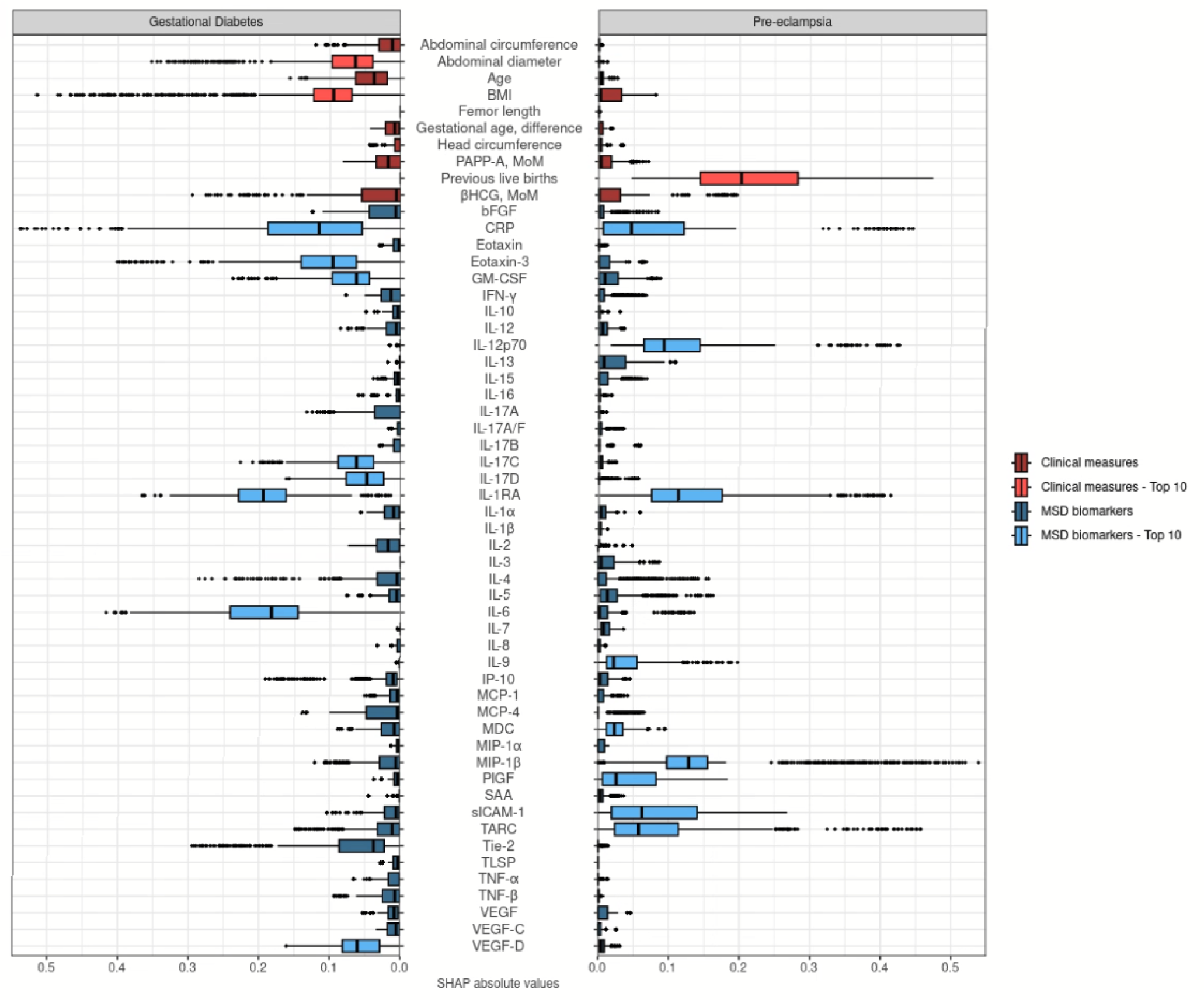
SHAP values for all prognostic features for gestational diabetes mellitus (GDM) and pre-eclampsia, respectively. The top ten features with the highest median value for each outcome are indicated with light colors. Only IL-1RA and CRP were part of both top ten lists. Features are colored based on the feature category as clinical measure (red) or MSD biomarker (blue). BMI, pre-pregnancy body mass index;MoM, multiple of medians; SHAP, Shapley additive explanation

Although the exploratory analysis yielded only a single probable biomarker for pre-eclampsia, our predictive modeling emphasized the combined significance of MIP-1β, IL-1RA, and IL-12p70. These biomarkers collectively had a greater impact than conventional risk factors such as age and BMI. Notably, the number of prior live births remained the most influential predictor (Figure 6B).

The remaining models had a low ability to discriminate and were not investigated further.

## Discussion

Here we present the biomarker profile of a large population-representative cohort of pregnant women around the 20th gestational week. We show that pregnancy has long-lasting effects and that the molecular level rewiring takes place at very early stages in the pregnancy, prior to the manifestation of complications at a clinically detectable level. Early changes in biomarkers were associated with development of obstetric outcomes up to 22 weeks later. Moreover, prior live births, complications from prior births, and smoking affect circulating biomarker levels, which may have a direct effect on complications and fetal growth. Machine learning models highlighted the joint significance of inflammatory markers, albeit the performance is not yet adequate for clinical deployment.

PREGCO is a prospective cohort recruited at a single hospital. Enrollment was high, above 61%. The cohort was generally comparable to other pregnancies in Denmark in the same time period. The difference in the frequency of pregnancy loss can most likely be attributed to an underreporting in the Danish National Patient Registry. The cohort was enrolled during a time of lowered activity in Denmark due to restrictions on both professional and private social interactions, hence the COVID-19 prevalence was therefore low at this time. Furthermore, enrollment took place when only the alpha COVID-19 variant was present, and later variants may have different effects on the immune system^39,40^. Furthermore, the testing regime and willingness to seek medical attention could have been different due to the COVID-19 pandemic. Relative to the sample size, the number of biomarkers investigated was large and some were highly correlated. We pruned highly correlated markers and utilized Bayesian models with conservative priors to provide regularization to the estimates to mitigate this and investigate issues of collinearity. We evaluated the generalization error of the machine learning models using a nested cross-validation approach, which yielded conservative estimates. However, we cannot rule out that some effects could not be detected due to the sample size, the large number of variables, and conservative Bayesian priors.

Pregnancy is a complex process that involves many changes in the body, including challenges to the immune system. These changes are necessary to allow the developing fetus to grow and thrive, but they can also increase the risk of obstetric complications. For instance, GM-CSF, a known regulator of fetal growth, was upregulated in pregnancies with a female child or smoking. The association between smoking and increased GM-CSF levels is hypothesized to be through an activation of the EGFR signaling pathway^28^. Furthermore, preterm birth or pre-eclampsia in a previous pregnancy was associated with lower levels of CRP. This is most likely medically induced due to preventive treatment with aspirin and progesterone, respectively.

The balance between pro- and anti-inflammatory cytokines is crucial for successful placentation. Pro-inflammatory cytokines, such as TNF-α, IL-1, and IL-6, play a role in the recruitment and activation of immune cells at the implantation site, which is essential for the establishment of a functional placenta^41^. IL-6 is involved in the regulation of angiogenesis, decidualization, and immune cell migration during early pregnancy, suggesting a crucial role for IL-6 in placentation^42^. Likewise, elevated levels of IL-17A have been associated with preterm labor and preterm premature rupture of membranes (PPROM)^43^. One proposed mechanism for this is IL-17A’s ability to promote pro-inflammatory cytokines and chemokines that can affect the development of the fetal-placental interface, especially together with TNF-α^43^. In our study, the pro-inflammatory cytokines of the IL-17-family were associated with disorders that have been linked with a negatively altered placentation, such as postpartum hemorrhage or preterm birth. The association of CRP, IL-13, IL-17B, and IL-17C with severe postpartum hemorrhage suggests that they may be involved in the regulation of hemostasis and low-grade inflammation early in pregnancy, leading to later obstetric complications. CRP is an acute-phase reactant produced by the liver in response to inflammation, infection, and tissue damage. IL-13 is a Th-2 cell cytokine involved in the regulation of immune response and tissue repair, whereas IL-17B and IL-17C are members of the IL-17 family of cytokines that are involved in the regulation of inflammation and immunity. Alterations in these biomarkers may affect the normal balance between pro- and anti-inflammatory factors and disrupt the physiological processes during and after delivery, leading for example to an increased risk of postpartum hemorrhage. More importantly, the biomarkers showed these associations to the outcome around gestational week 20, making them promising candidates for clinical use in early detection and prevention.

GDM is associated with increased risk of maternal and fetal morbidity and mortality, as well as an increased risk of developing type 2 diabetes later in life. The precise mechanisms underlying the development of GDM are not yet fully understood, but evidence suggests that a complex interplay of genetic and environmental factors is involved. In our study, Eotaxin-3, interleukin 1 receptor antagonist (IL-1RA), and IL-17D were associated with GDM. All three cytokines have been implicated in the regulation of immune responses, inflammation, and metabolic regulation. For example, elevated levels of Eotaxin-3 have been observed in pregnant women with pre-existing diabetes^44^. Antagonizing the pro-inflammatory cytokine IL-1, IL-1RA is an anti-inflammatory cytokine^45^. The interplay of IL-1 and IL-1RA and their dysregulation appear to be related to type 2 diabetes and GDM^32,46^. However, study results are diverging, as for example one study has shown that the IL-1RA levels in GDM patients are considerably lower than those of controls^32^, whereas another biomarker study showed inconsistence with our results, that elevated IL-1RA are associated with GDM^35^. In summary, our data show that the levels of these cytokines are altered in women developing GDM later in pregnancy and that the balance of pro- and antiinflammatory cytokines is necessary to maintain a normal glucose metabolism throughout pregnancy.

Additionally, the study highlights the importance of considering multiple biomarkers in assessing risk of obstetric complications, as each biomarker was associated with only one or a few of the outcomes. This suggests that a combination of biomarkers may be necessary to accurately identify women who are at an increased risk of developing complications, supported by the machine learning models.

Once they arise, many obstetric complications cannot be reversed or treated. Perinatal medicine therefore aims to identify high-risk populations early on, to ideally use therapies to reduce unfavorable maternal and fetal outcomes^47^. Dietary adaptations and insulin for GDM, antepartum fetal monitoring for stillbirth, aspirin for pre-eclampsia, and progesterone for preterm delivery are a few examples of such therapies that have been proposed in high-risk populations^48–53^. As such, it has been proposed that the typical care pyramid should be reversed, with the primary attention shifting to the early rather than later stages of pregnancy^47^. The presented biomarkers and biomarker combinations can potentially be used to further improve this pyramid of care by providing high-quality care to the patients at risk.

## Data Availability

The participants of this study did not give written consent for their data to be shared publicly, so due to the sensitive nature of the research supporting data is not available.

## Acknowledgements

The study was supported by funding from the Novo Nordisk Foundation (grant agreements NNF14CC0001 and NNF17OC0027594). The work was also carried out as a part of the BRIDGE Translational Excellence Programme (bridge.ku.dk) at the Faculty of Health and Medical Sciences, University of Copenhagen, funded by the Novo Nordisk Foundation (NNF18SA0034956).

## Competing Interest Statement

SB has ownerships in Hoba Therapeutics Aps, Novo Nordisk A/S, Lundbeck A/S, and managing board memberships in Proscion A/S. HSN received personal payment or honoraria for lectures and presentations from Ferring Pharmaceuticals, Merck, Astra Zeneca, Cook Medical, and Ibsa Nordic.

## Supplementary Information

### Meso Scale Diagnostics data normalization

We measured 47 inflammatory markers using the V-PLEX Human Biomarker 54-Plex Kit from Meso Scale Diagnostics (MSD). Due to low-quality assay validation, we excluded all measurements from the Th17 panel, resulting in 47 assays on six panels to be included in the analysis.

To address the batch effect exerted by the individual 96-well plate setup, we explored the corrective effects of three data pre-processing methods and four normalization methods with the aim of removing batch effects at panel level.

The MSD V-PLEX kit is based on electrochemiluminescence technology, where light emission from SULFO-TAG labels is measured as light intensity (“signal”). The signal values are within the dynamic range of the assay linearly associated with the concentration of the measured target. While this principle is used for concentration determination by a measured standard curve, we here used the signals directly to avoid any noise introduced by the measurement of a standard curve.

Due to the multiplex setup, plate-based batch effects were assessed by panel, as technical variation was assumed to be equal across the four to ten assays measured per panel. Our first aim was to remove any observed variation based on the 16 plates the samples were run on. We compared the combination of three pre-processing methods: *a*) log2-transformation, *b*) method *a* plus removal of outliers based on principal component analysis (PCA)^54^, and *c*) method *b* plus ComBat batch correction^55^, and four normalization methods: *i*) no normalization, *ii*) median normalization^56^, *iii*) quantile normalization^57^, and *iv)* MA normalization^58^. To assess the effectiveness of normalization, we performed a visual inspection of PCA plots, density plots, and box plots.

Based on visual inspection of the PCA plots of the log2-transformed data, we defined individual outlier limits for the six panels removing samples that exceeded these limits. Consequently, we removed 17 outliers on Angio1, 13 outliers on Chem1, 11 outliers on Cyto1, 41 outliers on Cyto2, 8 outliers on Pro1, and 9 outliers on Vascu2 (see limits on Supplementary Figure 2).

We found median normalization to sufficiently remove all batch effects observed at PC1 and PC2 and yielded overlapping curves with normal or near-normal distributions for the individual assays observed on density and box plots. Similar results were found for MA normalization, while the other methods yielded varying poorer results.

### Similarity reduction of markers using Hobohm II

Hierarchical clustering showed high similarity between markers, indicating redundancy in the data set. To address this, we employed the Hobolm II algorithm as outlined in Hobohm *et al*, 1992^20^. We used Spearman correlation as the correlation metric and tested cut-off levels at 0.3, 0.4, 0.5, 0.6, 0.7, 0.8, 0.9, and 1 (Supplementary Figure 3). We chose a cut-off of 0.5, resulting in 41 markers. Using this cut-off, we did not observe issues of collinearity in any of the statistical models.

### Bayesian regression models

For all statistical analyses we employed Bayesian regression models. All models were fit using rstanarm or brms^24,59,60^. Unless otherwise specified, all models were run for 10,000 iterations (5,000 warm-up and 5,000 sampling) with default settings for the sampler. Convergence was assessed by calculating r-hat statistics, looking for divergences and making sure the sample did not exceed the maximum tree depth after warm-up. A model had converged if and only if (1) all r-hat values < 1.01, (2) no divergences, and (3) no iterations exceeding the maximum tree depth. All parameters were centered prior to model fitting and MSD inflammatory markers were standardized so that the interpretation follows changes in standard deviations. Furthermore, we also inspected correlations of the posterior distributions to identify issues of collinearity.

### Bayesian robust linear regression

The Bayesian robust linear regression extends the classical ordinary least squares by assuming a Student t-distribution, thereby accommodating outliers. The degree of freedom is estimated directly from the data. Formally,

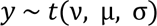

in which µ is the sum over the intercept and covariates,

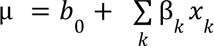

To complete the model, we specify a set of priors across the parameters in the model,

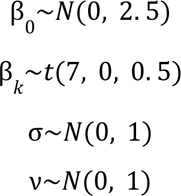

Posterior model checking was done by simulating 100 draws from the posterior and comparing with the observed distribution.

### Cause-specific parametric proportional hazards models

For analyzing the duration, or time to failure, we employed cause-specific parametric proportional hazards survival models, as implemented in rstanarm^59^. This takes into account that some women were lost to follow-up and are thus censored, and that there may be competing outcomes (e.g. induced labor, acute cesarean section, or scheduled cesarean section is a competing outcome to spontaneous vaginal birth). We compared two baseline hazards (cubic B-spline and M-spline) for each outcome and visually inspected the estimated baseline hazard curve, versus the observed curve. Model priors follow default settings, except the regression coefficients (β*_k_*) which were assigned a more conservative prior,

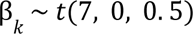

The prior induces regularization by forcing coefficients towards zero, i.e. a null effect.

### Birthweight to gestational age ratio

For the birth weight to gestational age ratio, the values were standardized according to the mean and standard deviation estimated from the Danish Medical Birth Registry (DMBR), including all births in 2020 (Supplementary Table 4). From the posterior, we defined a transformation,

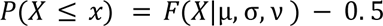

in which X is the estimated value of the marker, and F is the Student-t cumulative density distribution parametrized by the mean, standard deviation, and degrees of freedom estimated from the DMBR. The parameters were estimated using a Bayesian robust linear regression. The resulting value represents the change from the 50th percentile for a one-unit change in the variable.

### Supplementary Figures

**Supplementary Figure 1:**
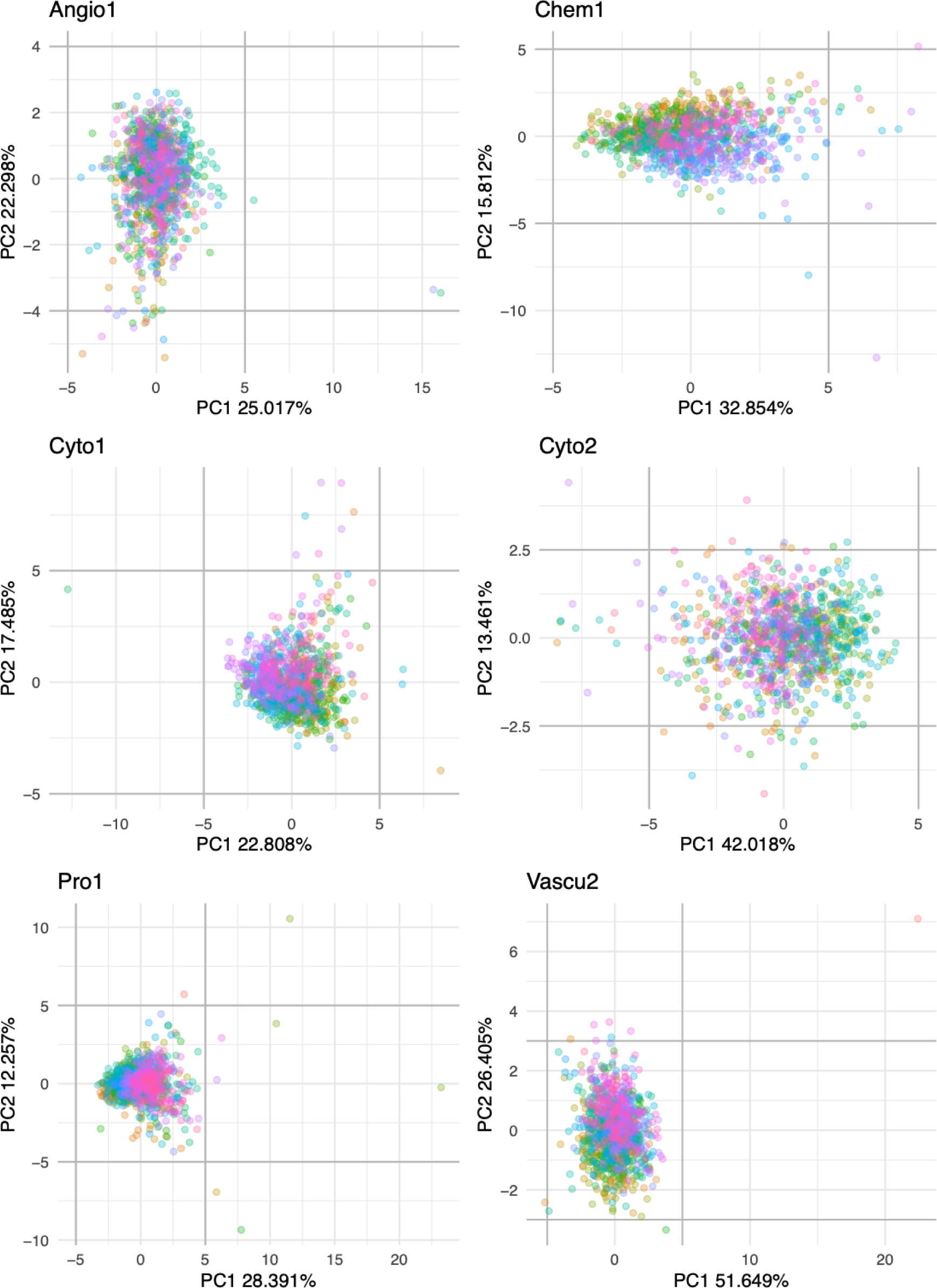
PCA plots for log2-transformed signal values colored by plate for the six MSD panels. Grey lines indicate selected limits for outlier detection.

**Supplementary Figure 2:**
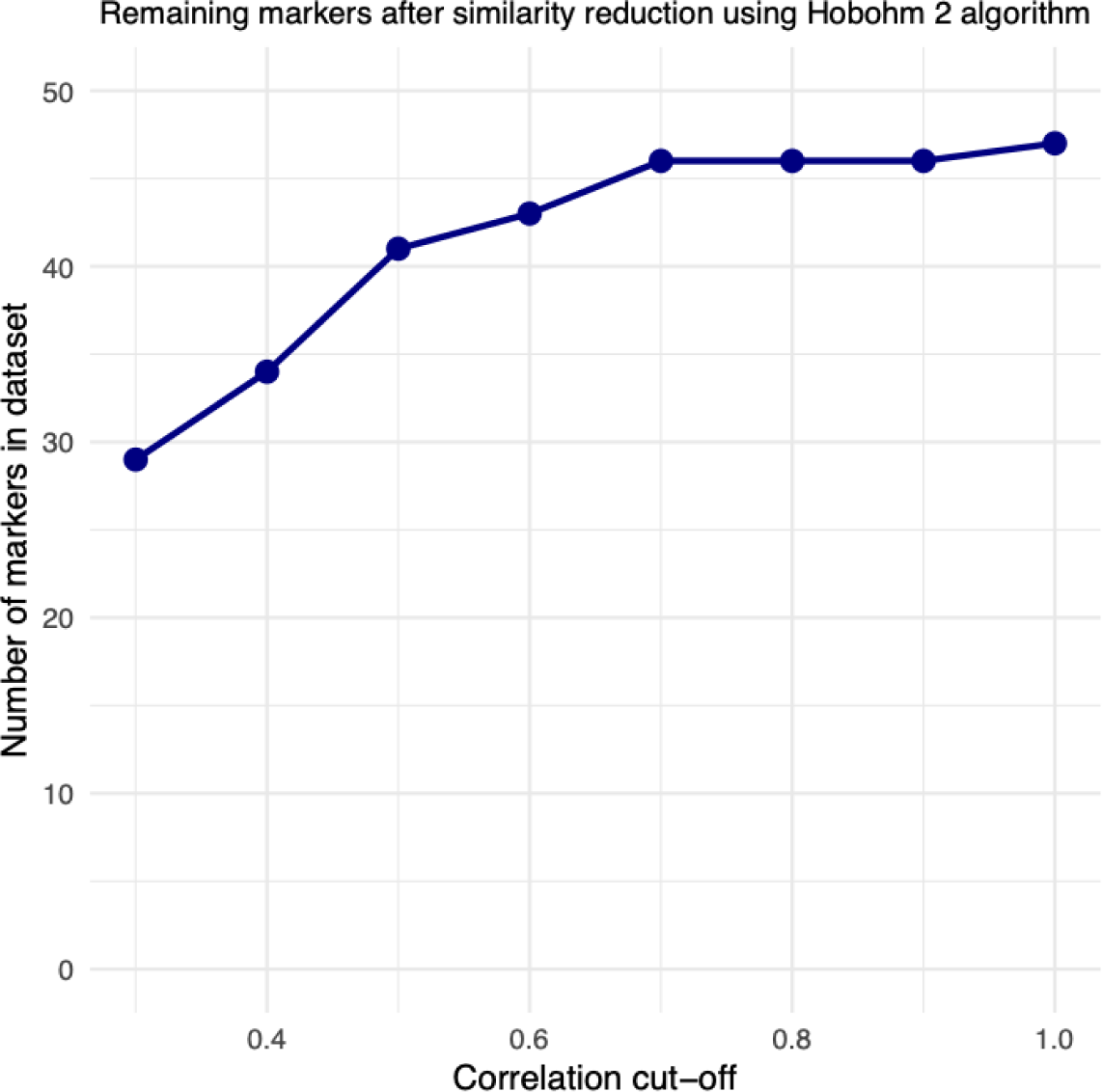
Number of markers remaining after pruning by the Hobohm II algorithm according to eight cut-off levels for the Spearman correlation coefficient.

**Supplementary Figure 3:**
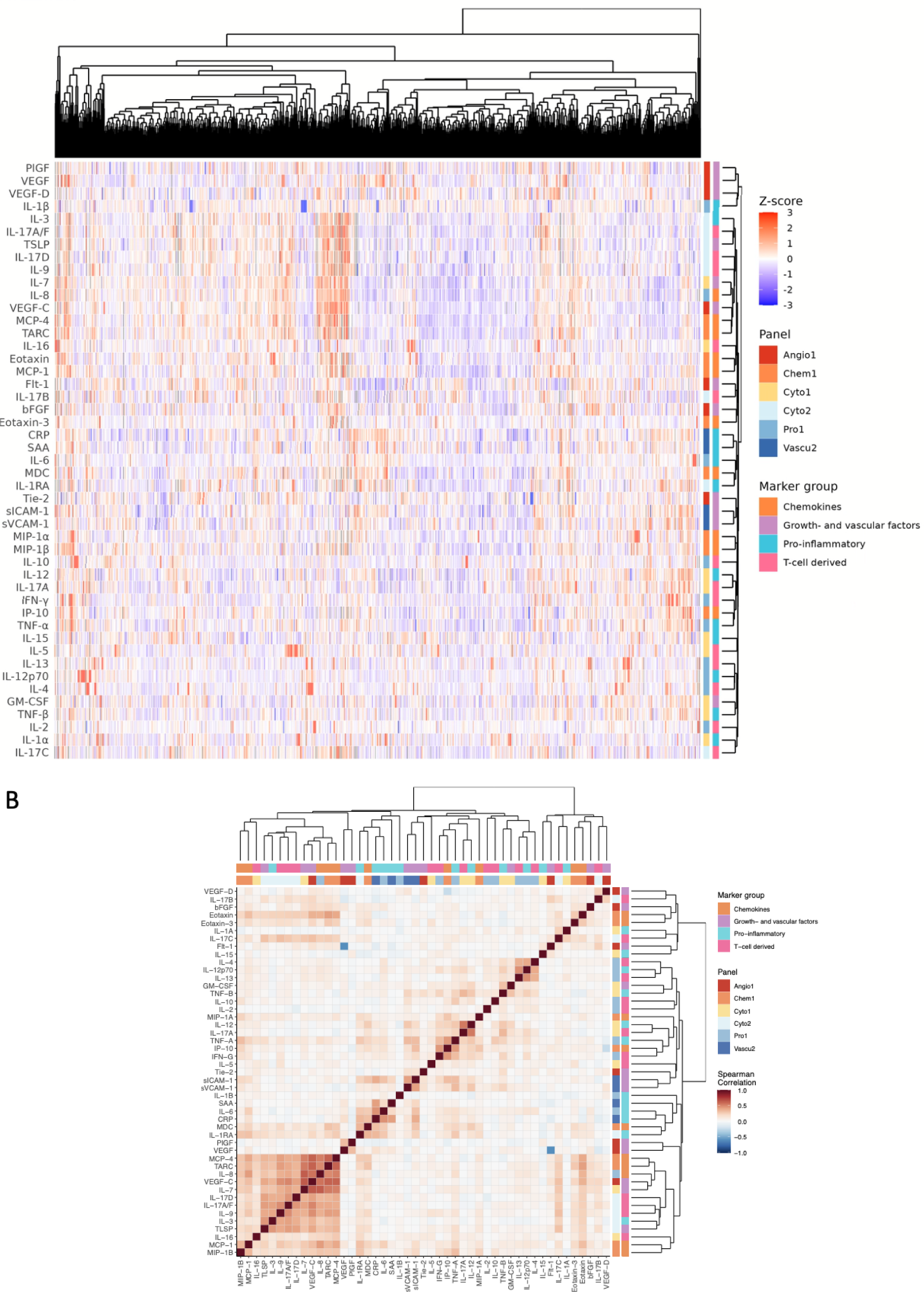
(a) Heatmap of women x markers, (b) Heatmaps of markers x markers.

### Supplementary Tables

**Supplementary Table 1:** Outcome descriptions, definitions, adjustments and exclusion criteria.

| **Outcome** | **Definition** | **Outcome-specific adjustments** | **Exclusion criteria** |
| --- | --- | --- | --- |
| Pre-eclampsia | Blood Pressure > 140 mmHg systolic and/or 90 mmHg diastolic & proteinuria | Previous number of live births,<br>Smoking,<br>PAPP-A MoM,<br>βHCG MoM,<br>ART (divided into IUI or IVF),<br>Ethnicity | Pre-gestational hypertension<br>Gestational hypertension diagnosed before inclusion<br>pre-eclampsia in previous pregnancy |
| Spontaneous vaginal birth | Birth not assisted by cesarean section or induction | Smoking,<br>Hemorrhage in early pregnancy,<br>UTI in current pregnancy,<br>Conisatio | n.a. |
| Gestational age at birth | Gestational age calculated from ultrasound. | Smoking,<br>Hemorrhage in early pregnancy,<br>UTI in current pregnancy,<br>Conisatio | n.a. |
| Preterm birth | Gestational age < 37+0 | Smoking,<br>Hemorrhage in early pregnancy,<br>UTI in current pregnancy,<br>Conisatio |  |
| Gestational diabetes mellitus (GDM) | Based on a two-hour 75 gram oral glucose tolerance test (OGTT); GDM with glucose ≥ 9.0 mmol/l in capillary whole blood or venous plasma | PCOS,<br>GDM in prior pregnancy | GDM diagnosed before inclusion<br>Diabetes Mellitus |
| Birth weight / gestational age ratio | Birth weight recorded immediately after birth by midwife or doctor. |  |  |
| Severe Postpartum hemorrhage | 1000 mL blood loss or more within 24 hours | ART (divided into IUI or IVF),<br>Hemorrhage in early pregnancy,<br>Cesarean section in prior pregnancy |  |
| Acute cesarean section | Delivery by acute cesarean section, | Cesarean section in prior pregnancy |  |

| any degree |  |  |  |
| --- | --- | --- | --- |
| Any complication | Pre-eclampsia, GDM, Severe PPH, Preterm birth, or acute cesarean section. | Previous number of live births, Smoking, PAPP-A MoM, βHCG MoM, ART (divided into IUI or IVF), PE in prior pregnancy, GDM in prior pregnancy, PCOS, UTI in current pregnancy, Cesarean section in prior pregnancy, Conisatio, Ethnicity | Pre-gestational hypertension<br>Gestational hypertension diagnosed before inclusion<br>pre-eclampsia in previous pregnancy<br>GDM diagnosed before inclusion<br>Diabetes Mellitus (type 1 or 2) |

Abbreviations: ART, assisted reproductive technology; GDM, gestational diabetes mellitus; IUI, intrauterine insemination; IVF, in vitro fertilization; MoM, multiple of the median; n.a., not applicable; OGTT, oral glucose tolerance test; PCOS, polycystic ovary syndrome; PE, Pre-eclampsia; PPH, postpartum hemorrhage; UTI, urinary tract infection

**Supplementary Table 2:**
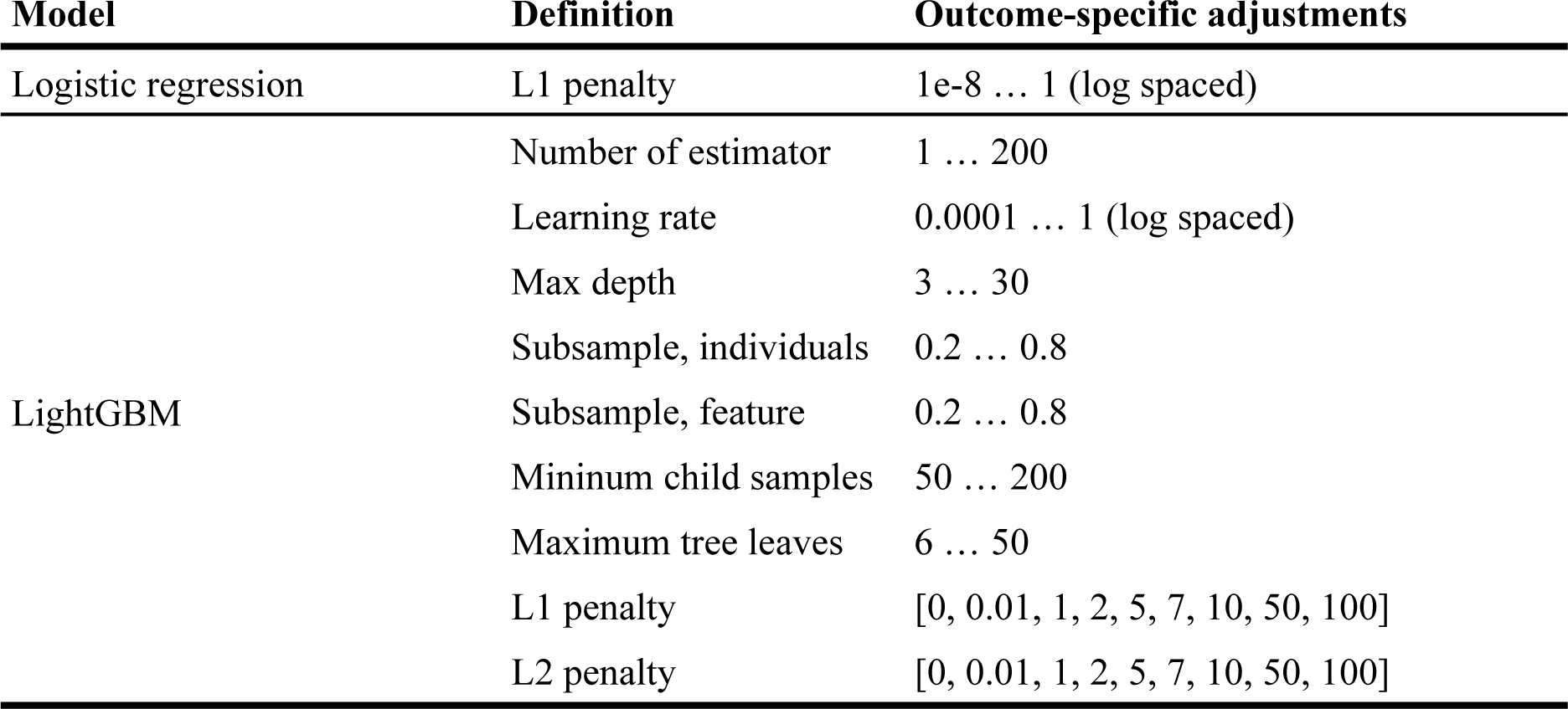
Hyperparameter values.

**Supplementary Table 3:** Comparison of PREGCO and the Danish Medical Birth Registry (DMBR).

| Variable |  | PREGCO<br>(n=1,049) | DMBR 2020<br>(n=60,573) | Difference |
| --- | --- | --- | --- | --- |
| Maternal age, years* |  | 31.7 (4.5) | 30.7 (4.7) | 1.0 (0.72; 1.27, p < 0.001) |
| Pre-pregnancy BMI, m/kg<sup>2</sup>* |  | 24.1 (4.8) | 24.8 (5.4) | -0.7 (-0.99; -0.41, p < 0.001) |
| Smoking during pregnancy** |  | 40 (3.8%) | 4,660 (7.7%) | 0.49 (0.35; 0.67, p < 0.001) |
| Parity** | 0 | 576 (54.9%) | 22,644 (37.3%) | p < 0.001 |
|  | 1 | 375 (35.7%) | 20,421 (33.7%) |  |
|  | 2 | 81 (7.7%) | 10,011 (16.5%) |  |
|  | 3+ | 17 (1.6%) | 7,507 (12.4%) |  |
| Number of Pregnancy Losses** | 0 | 810 (77.2%) | 52,145 (86%) | p < 0.001 |
|  | 1 | 194 (18.5%) | 6,965 (11.5%) |  |
|  | 2 | 29 (2.8%) | 1,230 (2.0%) |  |
|  | 3+ | 16 (1.5%) | 282 (0.5%) |  |
| Sex of child, male** |  | 521 (50.5%) | 30,983 (51%) | 0.96 (0.86; 1.07, p=0.44) |

\* Z-test; \*\* $\chi^2$ test

**Supplementary Table 4:** Birth weight-gestational duration ratio parameters estimated from the Danish Medical Birth Registry (DMBR).

| Parameter | Median | 95% bCI |
| --- | --- | --- |
| $\mu$ | 12.61 | 12.59-12.62 |
| $\sigma$ | 1.54 | 1.53-1.56 |
| $\nu$ | 6.88 | 6.54-7.24 |

**Supplementary Table 5:**
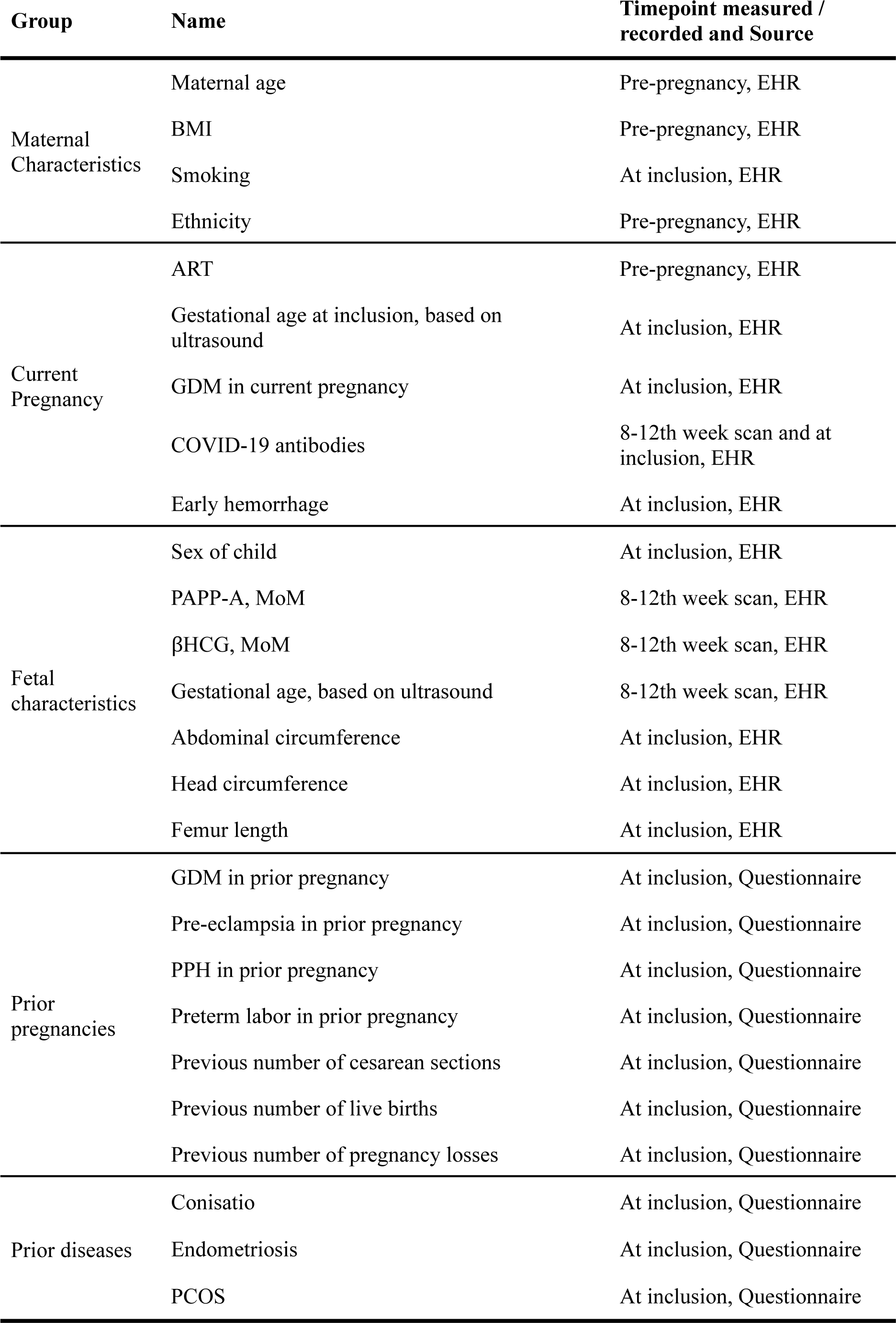
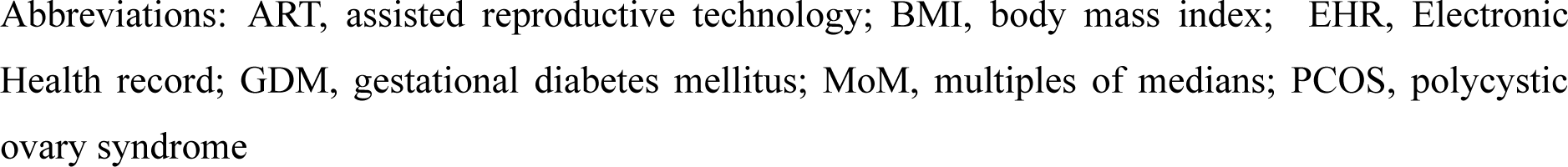
Variables and sources.

| Group | Name | Timepoint measured / recorded and Source |
| --- | --- | --- |
| Maternal Characteristics | Maternal age | Pre-pregnancy, EHR |
|  | BMI | Pre-pregnancy, EHR |
|  | Smoking | At inclusion, EHR |
|  | Ethnicity | Pre-pregnancy, EHR |
| Current Pregnancy | ART | Pre-pregnancy, EHR |
|  | Gestational age at inclusion, based on ultrasound | At inclusion, EHR |
|  | GDM in current pregnancy | At inclusion, EHR |
|  | COVID-19 antibodies | 8-12th week scan and at inclusion, EHR |
|  | Early hemorrhage | At inclusion, EHR |
| Fetal characteristics | Sex of child | At inclusion, EHR |
|  | PAPP-A, MoM | 8-12th week scan, EHR |
| | βHCG, MoM | 8-12th week scan, EHR |
|  | Gestational age, based on ultrasound | 8-12th week scan, EHR |
|  | Abdominal circumference | At inclusion, EHR |
|  | Head circumference | At inclusion, EHR |
|  | Femur length | At inclusion, EHR |
| Prior pregnancies | GDM in prior pregnancy | At inclusion, Questionnaire |
|  | Pre-eclampsia in prior pregnancy | At inclusion, Questionnaire |
|  | PPH in prior pregnancy | At inclusion, Questionnaire |
|  | Preterm labor in prior pregnancy | At inclusion, Questionnaire |
|  | Previous number of cesarean sections | At inclusion, Questionnaire |
|  | Previous number of live births | At inclusion, Questionnaire |
|  | Previous number of pregnancy losses | At inclusion, Questionnaire |
| Prior diseases | Conisatio | At inclusion, Questionnaire |
|  | Endometriosis | At inclusion, Questionnaire |
|  | PCOS | At inclusion, Questionnaire |

Abbreviations: ART, assisted reproductive technology; BMI, body mass index; EHR, Electronic Health record; GDM, gestational diabetes mellitus; MoM, multiples of medians; PCOS, polycystic ovary syndrome

**Supplementary Table 6:** Results from cross-validation of machine learning models. The highest ranking model for each outcome is highlighted in bold, based on the binary cross entropy from the inner CV.

| Outcome | Model | Development (inner CV) |  |  | Interval validation (outer CV) |  |
| --- | --- | --- | --- | --- | --- | --- |
|  |  | Binary Cross Entropy | ROC-AUC | AUPRC | ROC-AUC | AUPRC |
| Acute Sectio | **LASSO** | **0.216** | **0.638** | **0.130** | **0.590**<br>(0.515-0.667) | **0.086**<br>(0.057-0.126) |
|  | LightGBM | 0.216 |  |  |  |  |
| Any complication | **LASSO** | **0.599** | **0.630** | **0.457** | **0.615**<br>(0.576-0.655) | **0.421**<br>(0.367-0.477) |
|  | LightGBM | 0.602 |  |  |  |  |
| Gestational Diabetes | LASSO | 0.268 |  |  |  |  |
|  | **LightGBM** | **0.266** | **0.697** | **0.219** | **0.708**<br>(0.644-0.766) | **0.176**<br>(0.126-0.238) |
| Pre-eclampsia | LASSO | 0.150 |  |  |  |  |
|  | **LightGBM** | **0.149** | **0.655** | **0.125** | **0.672**<br>(0.580-0.758) | **0.073**<br>(0.042-0.115) |
| Preterm birth | LASSO | 0.172 |  |  |  |  |
|  | **LightGBM** | **0.171** | **0.610** | **0.118** | **0.564**<br>(0.460-0.66) | **0.071**<br>(0.041-0.132) |
| Severe PPH | LASSO | 0.349 |  |  |  |  |
|  | **LightGBM** | **0.349** | **0.628** | **0.198** | **0.587**<br>(0.530-0.638) | **0.152**<br>(0.117-0.193) |

## Notes

### Author Declarations

The PREGCO study was approved by the Knowledge Centre for Data Protection and Compliance, The Capital Region of Denmark (P-2020-255), and by the Scientific Ethics Committee of the Capital Region of Denmark (journal number H-20022647). All PREGCO participants provided written informed consent. Danish legislation allows for register-based research to be conducted without the consent of participants and without ethical committee approval. Registry data, i.e. the Danish Medical Birth Registry, was held at Statistics Denmark, which is the Danish national statistical institution.

